# The clinical feature of triple-negative breast cancer in Beijing, China

**DOI:** 10.1101/2021.08.03.21261573

**Authors:** Hui Zhao, Yueqing Feng, Junzheng Yang

## Abstract

**Objective:** To collect and analyze the clinical feature of triple-negative breast cancer (TNBC) in Beijing, to provide the information for the local oncologist to make sound treatment plans.

**Methods:** The clinical data of 280 breast cancer patients with TNBC admitted to the oncology hospital of China Academy of Medical Sciences were collected and divided into (recurrence and metastasis) group and non-(recurrence and metastasis) group. Breast cancer patients with TNBC were classified according to age distribution, family history of breast cancer, pathological type, histological grade, clinical stage, tumor thrombus, tumor size and lymph node metastasis. and 15 BRCA1 gene SNP loci were also measured by a high throughput Mass ARRAY time-of-flight mass spectrometry biochip system and compared the expression of 15 BRCA1 gene SNP loci between (recurrence and metastasis) group and non-(recurrence and metastasis) group. χ2 test was used to analyze the difference between two groups, and P<0.05 considered statistically significant.

**Results:** A total of 280 breast cancer patients with TNBC were enrolled in this study, median age 45 years old. There were 117 cases breast cancer patients with TNBC in (recurrence and metastasis) group, accounting for 41.79% in total breast cancer patients with TNBC and 163 cases breast cancer patients with TNBC in non-(recurrence and metastasis) group, accounting for 58.21% in total breast cancer patients with TNBC; There was no significant difference in age distribution, family history of breast cancer, pathological type and histological grade between non-(recurrence and metastasis) group and (recurrence and metastasis) group (P>0.05); but there were significant differences in clinical stage, vascular tumor thrombus, tumor size and lymph node metastasis between two groups (P<0.05); and then we compared the expression of 15 BRCA1 gene SNP loci in (recurrence and metastasis) group and non-(recurrence and metastasis) group, found that BRCA1gene rs 12516 CC loci (38.8% VS 44.4%), BRCA1gene rs 12516 TT loci (15.6% VS 10.4%), BRCA1 gene rs 16940 CC loci (15.1% VS 10.4%), BRCA1 gene rs 16940 TT loci (39.0% VS 44.8%), BRCA1 gene rs 16941 AA loci (38.1% VS 44%), BRCA1 gene rs 16941 GG loci (15.0% VS 10.3%), BRCA1 gene rs16942 AA loci (39.0% VS 44.8%), BRCA1 gene rs16942 GG loci (15.1% VS 10.4%), BRCA1gene rs799906 CC loci (15.9% VS 10.4%), BRCA1gene rs799906 TT loci (38.7% VS 44.8%), BRCA1gene rs799917 CC loci (38.7% VS 44.4%), BRCA1gene rs799917 TT loci (15.7% VS 10.4%), BRCA1gene rs1060915 CC loci (15.5% VS 10.4%), BRCA1gene rs1060915 TT loci (39.1%VS 44.8%), BRCA1gene rs1799966 AA loci (37.7% VS 44.4%), BRCA1gene rs1799966 GG loci (15.1% VS 10.4%), BRCA1 Gene rs2070833 AA loci (3.1% VS 7.0%), BRCA1 Gene rs2070833 CC loci (56.3% VS 51.3%), BRCA1gene rs3737559 GG loci(78.5% VS 84.5%), BRCA1gene rs3737559 GA loci(19.0% VS 14.6%), BRCA1gene rs8176199 AA loci (60.5% VS 64.6%), BRCA1gene rs8176318 GG loci (38.4% VS 43.4%), BRCA1gene rs8176318 TT loci (15.1% VS 10.6%), BRCA1gene rs8176323 CC loci (38.6% VS 43.9%), BRCA1gene rs8176323 GG loci (15.2% VS10.5%), BRCA1gene rs11655505 AA loci (14.9% VS 10.4%), BRCA1gene rs11655505 GG loci (39.1% VS 44.8%) had a difference at the accident rate between recurrence and metastasis group and non-(recurrence and metastasis) group, but the frequencies of genotypes in the (recurrence and metastasis) group and non-(recurrence and metastasis) group were similar, there was no statistical significant correlation between the SNP genotype of the BRCA1 gene and the recurrence and metastasis risk of TNBC (P>0.05).

**Conclusions:** There were higher recurrence and metastasis (41.79%) in total 280 cases breast cancer patients with TNBC in Beijing area; breast cancer patients with TNBC in Beijing area had a unique clinical feature no matter at clinical stage, vascular tumor thrombus, tumor size and lymph node metastasis or the expression of 15 BRCA1 gene SNP loci, those data may provide some information for clinical staff for TBNC treatment.

## 1 Introduction

Breast cancer is a kind of cancers with the higher incident rate in women in the world. The latest statistics showed that nearly 1.5 million women suffered from breast cancer every year, accounting for 25%-30% of all malignant tumors in women. Incidence rate of breast cancer has been increasing year by year with the improvement of living standard and lifestyle and the deterioration of living environment in China^[1]^. Breast cancer is a highly heterogeneous disease, and there are great differences in tissue morphology, molecular phenotypes, clinical manifestations, therapeutic effects and prognosis^[2]^.

Triple negative breast cancer (TNBC) is a kind of subtype of breast cancer with negative estrogen receptor (ER), negative progesterone receptor (PR) and negative human epidermal growth factor receptor 2 (HER2). accounting for 15∼20% in all breast cancer types^[3]^. It has the characteristics of poor tumor differentiation, strong invasiveness, easy local recurrence and distant metastasis^[4]^. The effective treatment methods are mainly chemotherapy and radiotherapy, and it is often insensitive to radiotherapy and chemotherapy. The present study found that TNBC is a group of tumors with different biological behavior. There are obvious differences in molecular biological characteristics, clinical manifestations and prognosis, and can be divided into more different subtypes^[5]^.

BRCA1 gene is located in human 17q21 with a length of about 100kb, including 24 exons and 22 introns, there are 22 exons with transcription and coding functions, and finally encode into phosphorylated protein containing 1863 amino acids, which plays a role in DNA damage repair, homologous recombination, cell cycle regulation, transcriptional activation and inhibition and centrosome replication^[6]^. BRCA1 gene is the most common susceptibility gene for breast cancer. The proportion of breast cancer with BRCA1 mutation in the lifetime is as high as 60%∼80%, and the onset age is more than 20 years earlier than that of the general population^[7]^. A large number of studies have shown that the proportion of TNBC in BRCA1 mutation carriers is higher, and often shows large size, poor differentiation, strong invasion, and easy to have local recurrence and distant metastasis; more than 30% of the TNBC are associated with mutations in the BRCA1 gene^[8]^.

Here, we investigated and analyzed the clinical features of TNBC patients in Beijing, compared the difference between (recurrence and metastasis) group and non-(recurrence and metastasis) group in age distribution, family history of breast cancer, pathological type, histological grade, clinical stage, vascular tumor thrombus, tumor size and lymph node metastasis and the expression of 15 BRCA1 gene SNP loci, which may provide some clues for clinic staff for TNBC treatment in Beijing.

## 2 Investigation subject

The investigation subjects in this study were TNBC breast cancer patients who were admitted to the Cancer Hospital of China Academy of Medical Sciences in the past 5 years. There were a total of 280 breast cancer patients, They were Han population of China, had complete demographic data, clinical pathological data and follow-up data. The patients in the study were divided into (recurrence and metastasis) group and non-(recurrence and metastasis) group according to whether there was recurrence or metastasis. Usually according to the criterion, patients who had no local recurrence or distant metastasis for more than 3 years from diagnosis to enrollment were considered there was no recurrence and metastasis.

All the patients in the study were diagnosed as TNBC patients, when expression of ER, PR and HER2 were negative considered as TNBC patients. The expression of ER and PR was determined by immunohistochemistry (IHC), The expression of HER-2 was determined by IHC or fluorescence in situ hybridization (FISH). When the positive ratio <10% in breast cancer cells considered ER negative and PR negative, the criteria for HER2 negative were immunohistochemical staining (-)/(+) or fluorescence in situ hybridization without amplification.

All enrolled patients were followed up for a long time by consulting the original medical records, outpatient reexamination and telephone follow-up, and the tumor recurrence, metastasis and survival of enrolled patients were recorded in detail.

## 3 Investigation methods

TNBC breast cancer patients were classified according to age distribution, family history of breast cancer, pathological type, histological grade, clinical stage, vascular tumor thrombus, tumor size and lymph node metastasis and the expression of 15 BRCA1 gene SNP loci, and compared the difference between (recurrence and metastasis) group and non-(recurrence and metastasis) group.

For BRCA1 gene SNP determination, firstly BRCA1 gene was retrieved from the SNP public database provided and the human genome genetic variation integration map published by the thousand people genome project (http://hapmap.ncbi.nlm.nih.gov/), all SNP loci related to the known BRCA1 gene information in the Chinese Han population were selected, combined with the statistical characteristics of minimum allele frequency (MAF) and Hardy Weinberg equilibrium *p* value (HWPval), the above candidate SNP sites were screened to obtain the best labeled SNP. Selection requirements was MAF>0.1 and HWPval>0.1. Then the labeled SNP sites were replaced and supplemented according to the location of SNP sites (missense mutation sites in exon region, UTR region, promoter region and intron region were usually selected, and synonymous mutation was not selected) and the positive SNP sites related to BRCA1 gene reported in the literature at home and abroad. Finally, sequence primers were designed for the selected loci by software. Considered the interaction between different primers, 15 SNP loci were selected finally. The specific information of SNP loci of BRCA1 gene is shown in Table 1.

**Table 1.** The information of 15 SNP loci of BRCA 1

| SNP loci | Functional location | Allele gene |
| --- | --- | --- |
| rs16940 | exon-synon | C/T |
| rs16941 | exon | A/G |
| rs16942 | exon | A/G |
| rs12516 | 3' UTR | C/T |
| rs799906 | promoter | C/T |
| rs1799966 | exon | A/G |
| rs799917 | exon-missense | C/T |
| rs3737559 | intron | A/G |
| rs1060915 | exon-synon | C/T |
| rs2070833 | intron | C/A |
| rs4793204 | promoter | A/G |
| rs8176199 | intron | C/A |
| rs8176318 | 3' UTR | G/T |
| rs8176323 | 3' UTR | C/G |
| rs11655505 | promoter | A/G |

## 4 Statistics

All data were analyzed by SPSS17.0 software. χ2 test were used for analyzing the difference between groups, P <0.05 considered as statistical significance.

## 5 Results

A total of 280 TNBC breast cancer patients were enrolled in this study, median age were 45 years old (24∼78 years old), there were 85 cases TNBC patients aged under 40 years old, and 195 cases TNBC patients aged above 40 years old, accounting for 30.36% and 69.64% in total TNBC breast cancer patients respectively. There were 117 cases TNBC patients occurred recurrence and metastasis, accounting for 41.79% in total 280 cases breast cancer patients with TNBC in Beijing area, There was no significant difference in age distribution, family history of breast cancer, pathological type and histological grade between relapse and metastasis group and disease-free survival group (P>0.05). There were significant differences in clinical stage, vascular tumor thrombus, tumor size and lymph node metastasis (P<0.05). In the (recurrence and metastasis) group, 58 cases (49.6%) had late clinical stage, 62 cases (53.0%) had vascular tumor thrombus, 74 cases (63.2%) had large tumor and 84 cases (71.8%) had lymph node metastasis; the data were shown at table 3.

**Table 2.**
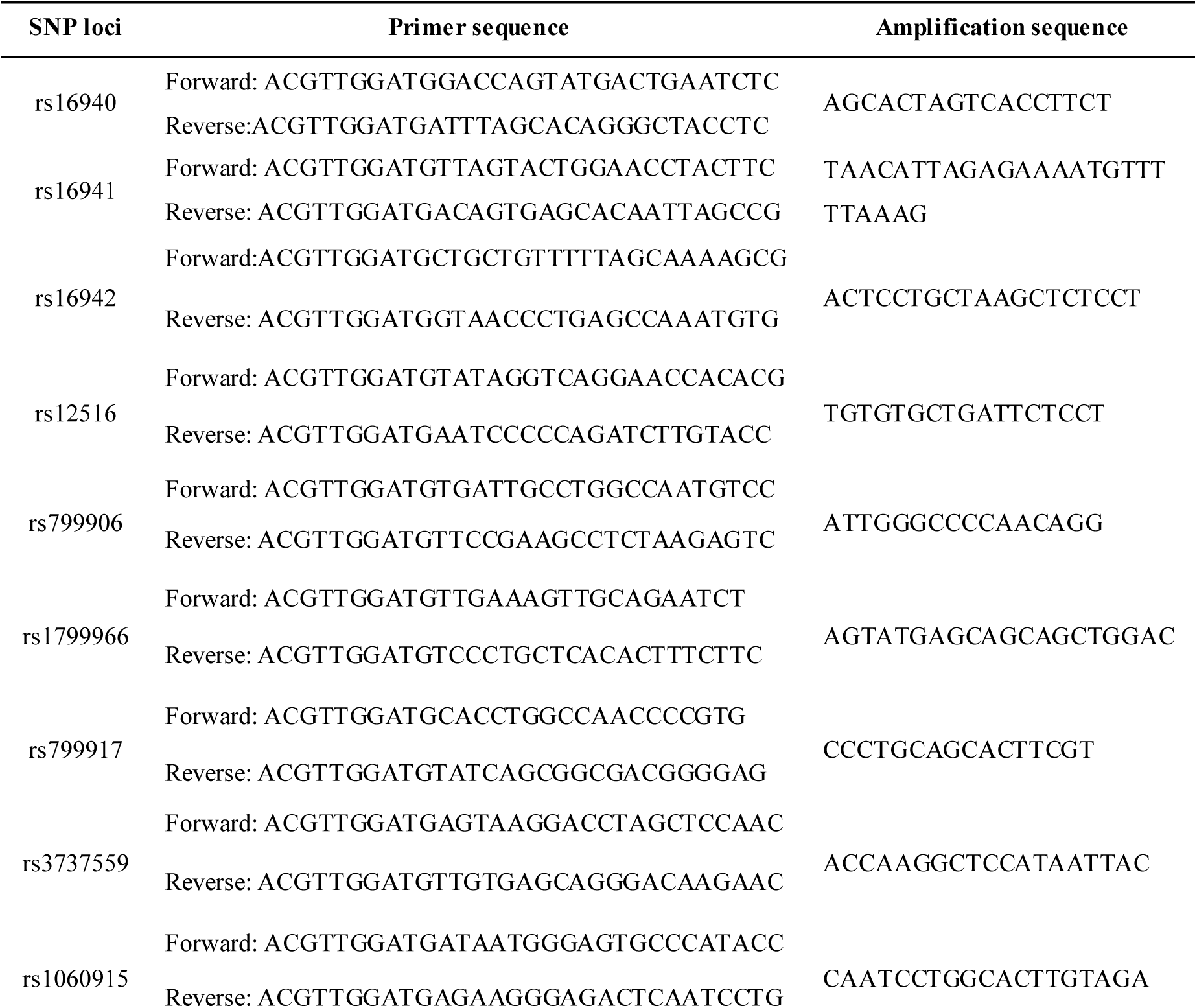
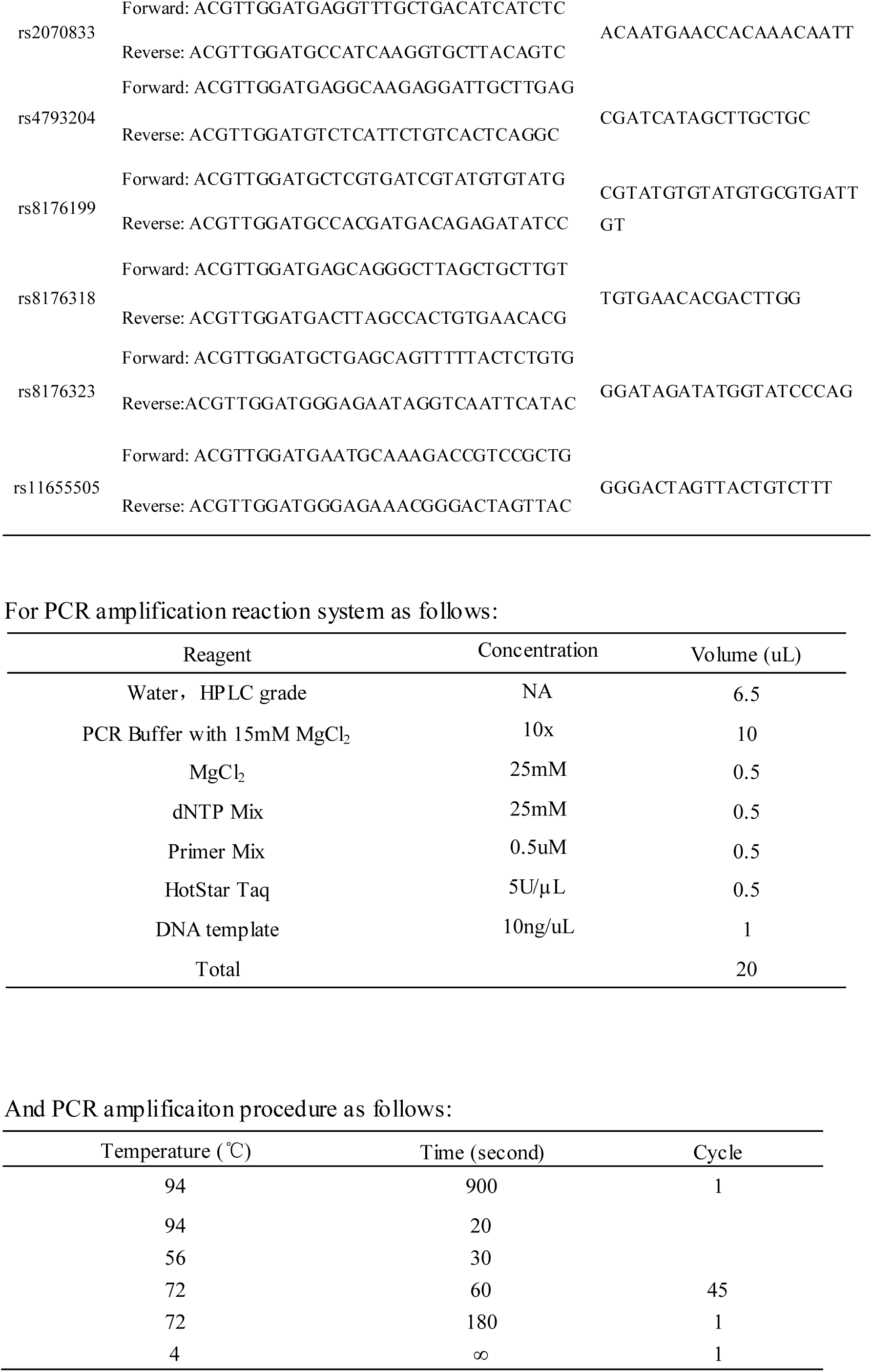
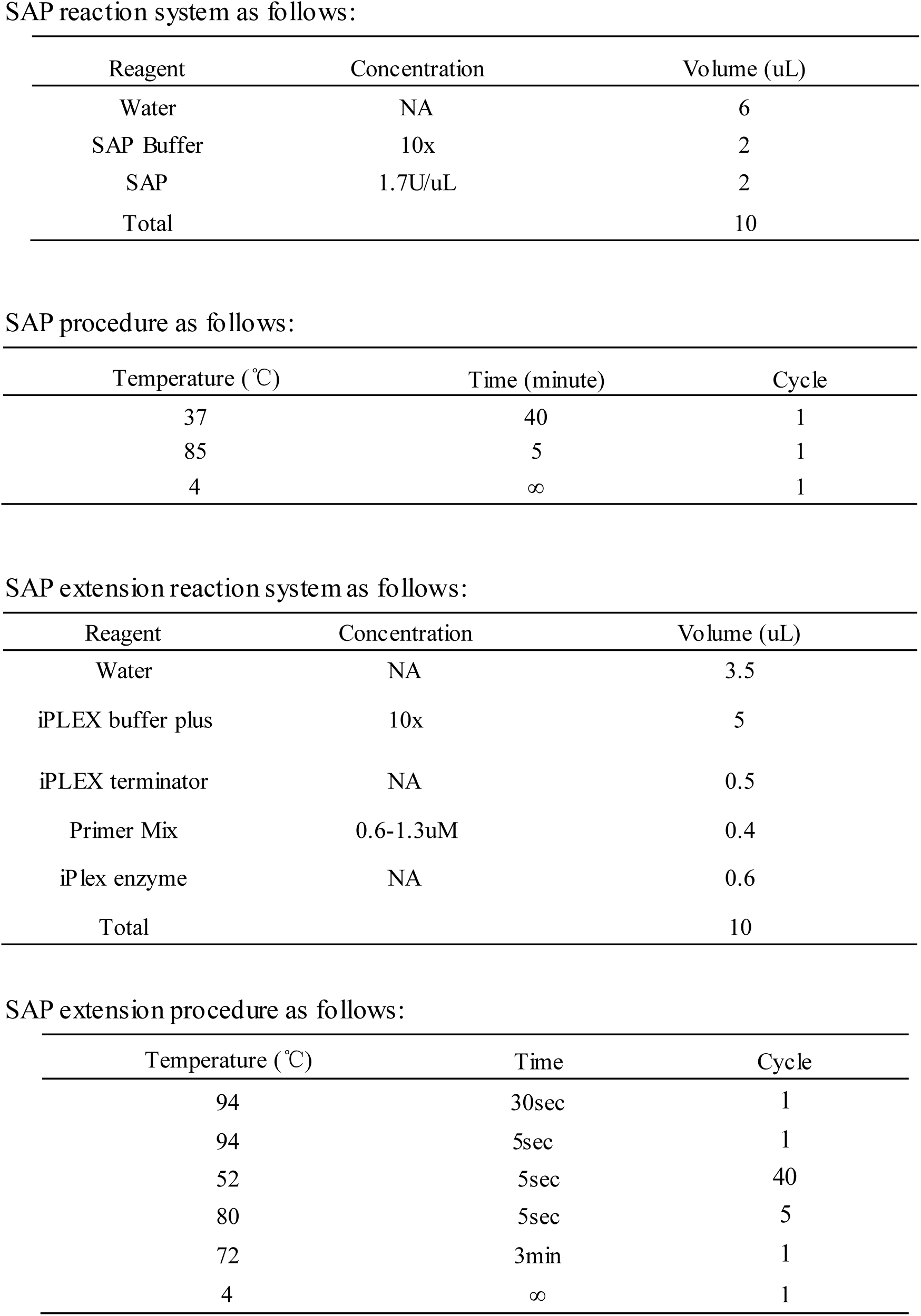
Primer sequences of 15 SNP loci of BRCA 1

**Table 3.** the demographic characteristics of TNBC Patients in the study

|  | non-(Recurrence and metastasis) group | (Recurrence and metastasis) group | P value |
| --- | --- | --- | --- |
|  | Cases (%) | Cases (%) |  |
| **Age** |  |  | 0.696 |
| ≤40 | 48 (29.4) | 37 (31.6) |  |
| >40 | 115 (70.6) | 80 (68.4) |  |
| **Family history** |  |  | 0.223 |
| No | 150 (92.0) | 105 (89.7) |  |
| Yes | 10 (6.1) | 12 (10.3) |  |
| Unknown | 3 (1.9) | 0 (0.0) |  |
| **Clinical stage** |  |  | **0.000** |
| I+II stage | 141 (86.5) | 54 (46.2) |  |
| III+IV stage | 19 (11.7) | 58 (49.6) |  |
| Unknown | 3 (1.8) | 5 (4.2) |  |
| **Pathological type** |  |  | 0.233 |
| Invasive nonspecific carcinoma | 131(80.4) | 102(87.2) |  |
| Other type | 29 (17.8) | 15(12.8) |  |
| Unknown | 3 (1.8) | 0 (0.0) |  |
| **Histological grade** |  |  | **0.111** |
| Level1+level 2 | 88 (54.0) | 56 (47.9) |  |
| Level 3 | 53 (32.5) | 51 (43.6) |  |
| Unknown | 22 (13.5) | 10 (8.5) |  |
| **Vascular tumor thrombus** |  |  | **0.017** |
| No | 98 (60.1) | 52 (44.4) |  |
| Yes | 65 (39.9) | 62(53.0) |  |
| Unknown | 0 (0.0) | 3 (2.6) |  |
| **Tumor size** |  |  | **0.004** |
| ≤2cm | 82 (50.3) | 39 (33.3) |  |
| >2cm | 75 (46.0) | 74 (63.2) |  |
| Unknown | 6 (3.7) | 4 (3.4) |  |
| **Lymph node metastasis** |  |  | **0.000** |
| No | 112 (68.7) | 28 (23.9) |  |
| Yes | 48 (29.5) | 84 (71.8) |  |
| Unknown | 3 (1.8) | 5 (4.3) |  |

And then we detected and analyzed the expression of 15 BRCA1 gene SNP loci in (recurrence and metastasis) group and non-(recurrence and metastasis) group, found that BRCA1gene rs 12516 CC loci (38.8% VS 44.4%), BRCA1gene rs 12516 TT loci (15.6% VS 10.4%), BRCA1 gene rs 16940 CC loci (15.1% VS 10.4%), BRCA1 gene rs 16940 TT loci (39.0% VS 44.8%), BRCA1 gene rs 16941 AA loci (38.1% VS 44%), BRCA1 gene rs 16941 GG loci (15.0% VS 10.3%), BRCA1 gene rs16942 AA loci (39.0% VS 44.8%), BRCA1 gene rs16942 GG loci (15.1% VS 10.4%), BRCA1gene rs799906 CC loci (15.9% VS 10.4%), BRCA1gene rs799906 TT loci (38.7% VS 44.8%), BRCA1gene rs799917 CC loci (38.7% VS 44.4%), BRCA1gene rs799917 TT loci (15.7% VS 10.4%), BRCA1gene rs1060915 CC loci (15.5% VS 10.4%), BRCA1gene rs1060915 TT loci (39.1% VS 44.8%), BRCA1gene rs1799966 AA loci (37.7% VS 44.4%), BRCA1gene rs1799966 GG loci (15.1% VS 10.4%), BRCA1 Gene rs2070833 AA loci (3.1% VS 7.0%), BRCA1 Gene rs2070833 CC loci (56.3% VS 51.3%), BRCA1gene rs3737559 GG loci (78.5% VS 84.5%), BRCA1gene rs3737559 GA loci (19.0% VS14.6%), BRCA1gene rs8176199 AA loci (60.5%VS 64.6%), BRCA1gene rs8176318 GG loci (38.4% VS 43.4%), BRCA1gene rs8176318 TT loci (15.1% VS 10.6%), BRCA1gene rs8176323 CC loci (38.6% VS 43.9%), BRCA1gene rs 8176323 GG loci (15.2% VS 10.5%), BRCA1gene rs11655505 AA loci (14.9% VS 10.4%), BRCA1gene rs11655505 GG loci (39.1% VS 44.8%) had a difference at the accident rate between (recurrence and metastasis) group and non-(recurrence and metastasis) group, but the frequencies of genotypes in the (recurrence and metastasis) group and non-(recurrence and metastasis) group were similar, there was no statistical significant correlation between the SNP genotype of the BRCA1 gene and the recurrence and metastasis risk of TNBC (P>0.05), the results were shown at table 4.

**Table 4.** the expression of 15 BRCA1 gene SNP loci in TNBC patients

| SNP |  | Group A<br>Cases (%) | Group B<br>Cases(%) | P value |
| --- | --- | --- | --- | --- |
| BRCA1 gene | rs 12516 |  |  | 0.396 |
|  | CC | 62 (38.8) | 51(44.4) |  |
|  | TT | 25 (15.6) | 12(10.4) |  |
|  | TC | 73 (45.6) | 52(45.2) |  |
| BRCA1 gene | rs 16940 |  |  | 0.422 |
|  | CC | 24 (15.1) | 12(10.4) |  |
|  | TT | 62 (39.0) | 52(44.8) |  |
|  | TC | 73 (45.9) | 52(44.8) |  |
| BRCA1 gene | rs 16941 |  |  | 0.427 |
|  | AA | 61 (38.1) | 51(44.0) |  |
|  | GG | 24 (15.0) | 12 (10.3) |  |
|  | AG | 75 (46.9) | 53 (45.7) |  |
| BRCA1 gene rs16942 |  |  |  | 0.422 |
|  | AA | 62 (39.0) | 52 (44.8) |  |
|  | GG | 24 (15.1) | 12 (10.4) |  |
|  | GA | 73 (45.9) | 52 (44.8) |  |
| BRCA1gene rs799906 |  |  |  | 0.334 |
|  | CC | 26 (15.9) | 12 (10.4) |  |
|  | TT | 63 (38.7) | 52 (44.8) |  |
|  | TC | 74 (45.4) | 52 (44.8) |  |
| BRCA1gene rs799917 |  |  |  | 0.373 |
|  | CC | 61(38.4) | 51(44.4) |  |
|  | TT | 25 (15.7) | 12(10.4) |  |
|  | TC | 73 (45.9) | 52(45.2) |  |
| BRCA1gene rs1060915 |  |  |  | 0.389 |
|  | CC | 25 (15.5) | 12 (10.4) |  |
|  | TT | 63 (39.1) | 52(44.8) |  |
|  | TC | 73 (45.4) | 52 (44.8) |  |
| BRCA1gene rs1799966 |  |  |  | 0.391 |
|  | AA | 60 (37.7) | 51 (44.4) |  |
|  | GG | 24 (15.1) | 12 (10.4) |  |
|  | GA | 75 (47.2) | 52 (45.2) |  |
| BRCA1gene rs2070833 |  |  |  | 0.301 |
|  | AA | 5 (3.1) | 8 (7.0) |  |
|  | CC | 90 (56.3) | 59 (51.3) |  |
|  | CA | 65 (40.6) | 48 (41.7) |  |
| BRCA1gene rs3737559 |  |  |  | 0.348 |
|  | AA | 4 (2.5) | 1 (0.9) |  |
|  | GG | 128(78.5) | 98(84.5) |  |
|  | GA | 31(19.0) | 17(14.6) |  |
| BRCA1gene rs4793204 |  |  |  | 0.943 |
|  | GG | 115 (75.2) | 83 (74.8) |  |
|  | GA | 38 (24.8) | 28 (25.2) |  |
| BRCA1gene rs8176199 |  |  |  | 0.772 |
|  | AA | 98 (60.5) | 75 (64.6) |  |
|  | CC | 10 (6.2) | 6 ( 5.2) |  |
|  | CA | 54 (33.3) | 35 (30.2) |  |
| BRCA1gene rs8176318 |  |  |  | 0.494 |
|  | GG | 61(38.4) | 49 (43.4) |  |
|  | TT | 24(15.1) | 12(10.6) |  |
|  | GT | 74(46.5) | 52(46.0) |  |
| BRCA1gene rs8176323 |  |  |  | 0.463 |
|  | CC | 61 (38.6) | 50 (43.9) |  |
|  | GG | 24 (15.2) | 12 (10.5) |  |
|  | GC | 73 (46.2) | 52 (45.6) |  |
| BRCA1 gene rs11655505 |  |  |  | 0.444 |
|  | AA | 24 (14.9) | 12 (10.4) |  |
|  | GG | 63 (39.1) | 52 (44.8) |  |
|  | AG | 74 (46.0) | 52 (44.8) |  |

Group A: non-(recurrence and metastasis) group; Group B: (recurrence and metastasis) group.

## 6 Discussion

TNBC has the poor prognosis and worse survival, endocrine therapy and targeted therapy is ineffective for TNBC treatment; the effective treatment methods are mainly chemotherapy and radiotherapy, but they are often insensitive to radiotherapy and chemotherapy^[9,10]^, therefore, it is of important clinical significance to understand the clinical characteristics and pathogenesis of TNBC patients for local oncologist. In this study, we collected and analyzed the clinical data of a total 280 case of TNBC patients which admitted to the oncology hospital of China Academy of Medical Sciences. There were 85 cases TNBC patients aged under 40 years old, and 195 cases TNBC patients aged above 40 years old, accounting for 30.36% and 69.64% in total TNBC breast cancer patients respectively, patients there were 117 cases breast cancer patients occurred recurrence and metastasis, accounting for 41.79% in total 280 cases breast cancer patients with TNBC in Beijing area; clinical stage, vascular tumor thrombus, tumor size and lymph node metastasis in (recurrence and metastasis) group were significantly different from their in non-(recurrence and metastasis) group, those data demonstrated that the age of TNBC were 40 years old, and there were the higher recurrence and metastasis in TNBC patients in Beijing area.

And then we detected and analyzed expression of 15 BRCA1 gene SNP loci in (recurrence and metastasis) group and non-(recurrence and metastasis) group, found that there were many difference in several SNP loci, but the frequencies of genotypes in the (recurrence and metastasis) group and non-(recurrence and metastasis) group were similar, there was no statistical significant correlation between the SNP genotype of the BRCA1 gene and the recurrence and metastasis risk of TNBC (P>0.05). Those data demonstrated that the BRCA1 gene SNP loci in TNBC patients expression of BRCA1 gene in different TNBC patients, and the mechanism of TNBC was genetic variation, recurrence and metastasis of negative breast cancer is relatively complicated.

## Data Availability

The data used to support the findings of this study are available from the corresponding author upon request.

## 8 Acknowledgements

We would like to thank Mr. Ma Fei from the Department of oncology, cancer hospital, Chinese Academy of Medical Sciences for his guidance and help in this study.

